# An integrated deterministic-stochastic approach for forecasting the long-term trajectories of COVID-19

**DOI:** 10.1101/2020.05.13.20101303

**Authors:** Indrajit Ghosh, Tanujit Chakraborty

**Author notes:** Corresponding author’s.

## Abstract

The ongoing COVID-19 pandemic is one of the major health emergencies in decades that affected almost every country in the world. As of June 30, 2020, it has caused an outbreak with more than 10 million confirmed infections, and more than 500 thousand reported deaths globally. Due to the unavailability of an effective treatment (or vaccine) and insufficient evidence regarding the transmission mechanism of the epidemic, the world population is currently in a vulnerable position. The daily cases data sets of COVID-19 for profoundly affected countries represent a stochastic process comprised of deterministic and stochastic components. This study proposes an integrated deterministic-stochastic approach to forecast the long-term trajectories of the COVID-19 cases for Italy and Spain. The deterministic component of the daily-cases univariate time-series is assessed by an extended version of the SIR (SIRCX) model, whereas its stochastic component is modeled using an autoregressive (AR) time series model. The proposed integrated SIRCX-AR (ISA) approach based on two operationally distinct modeling paradigms utilizes the superiority of both the deterministic SIRCX and stochastic AR models to find the long-term trajectories of the epidemic curves. Experimental analysis based on the proposed ISA model shows significant improvement in the long-term forecasting of COVID-19 cases for Italy and Spain in comparison to the ODE-based SIRCX model.

## 1. Introduction

Coronavirus disease 2019 (COVID-19) is a rapidly spreading disease transmitted between people via respiratory droplets [1]. The COVID-19 pandemic is the most significant global crisis since the World War-II that affected almost all the countries throughout the world. The World Health Organization (WHO) declared COVID-19 a “global pandemic” on March 11, 2020. A notable characteristic of COVID-19 is its ability to cause unusually large case clusters via super-spreading in the absence of any specific antiviral treatment available to cure COVID [2]. The health consequences of the pandemic are devastating, and it has also triggered a global health concern. This has bought the scientific community to come up with different short-term and long-term forecasting models for a better understanding of the pandemic and mitigate the effects of this.

So far, various mathematical, statistical and machine learning models have been deployed to predict the disease dynamics and also to assess the efficiency of the intervention strategies in reducing the burden of COVID-19 [3, 4, 5, 6, 7, 8]. In previous studies, mathematical modeling approaches (e.g., SIR and SEIR models) [9, 10, 11] performed more effectively for predicting the long-term trajectories of the epidemic. In contrast, stochastic forecasting models (classical ARIMA and Wavelet-based forecasting model) [12] gave superior real-time short-term forecasts for some profoundly affected countries. However, a close look at the daily confirmed cases data sets of COVID-19 for different countries exhibits both the deterministic trend and stochastic behavior in the series. This indicates that the predictions of long-term trajectories of COVID-19 daily cases can be greatly improved by integrating both the deterministic and stochastic approaches. This study attempts to blend deterministic and stochastic methods for studying the long term trajectories of COVID-19 cases for Italy and Spain.

Motivated by the above discussion, we propose an integrated deterministic-stochastic model to describe the long-term trajectories of COVID-19 daily cases. The underlying characteristics of the epidemic curves show both the deterministic and stochastic nature that intrigued the need for developing an integrated deterministic-stochastic model to forecast the long-term trajectories of COVID-19 cases. In the first stage of the model, an extended version of the SIR model (SIRCX) is built that preserves the characteristic of the deterministic process in the long run. The deterministic SIRCX model leaves a certain part of the examined time series unexplained, and we overcome these uncertainties with the help of a stochastic model. In the second stage of the integrated approach, a stochastic autoregressive (AR) model is applied to analyze the residuals’ uncertain behavior produced by the compartmental SIRCX model. This newly introduced integrated SIRCX-AR model, we call it the ‘ISA model’, exploits the benefits of two methodologically contrasting paradigms and overcome their shortcomings. The proposed ISA model preserves both the long-term and short-term characteristics of the current pandemic time series when applied to the daily confirmed cases data sets of Italy and Spain. This combined approach is also useful for explaining complex autocorrelation structures in the COVID-19 time-series data. It also reduces the inductive bias and variances of the individual models from a modeler’s point of view. Besides that, we estimate the basic reproduction number, expected total cases after one month, and expected daily cases after two months. The proposed ISA model will be an excellent alternative for forecasting long-term trajectories of the epidemic curve for COVID-19 and other future pandemics.

The remainder of this paper is organized as follows. In Section 2, we discuss the data sets, data pre-processing steps, and the developments of the integrated deterministic-stochastic approach. In Section 3, the experimental findings are presented. Finally, the limitations of our methodology, along with discussions and future directions of the paper are given in Section 4.

## 2. Materials and methods

In this section, we describe the data sets and the necessary pre-processing stages before modeling the data. We further describe the formulation of the proposed integrated approach, along with the component models.

### 2.1. Data

We focus on the daily figures of confirmed cases for Italy and Spain. These data sets are retrieved by the Global Change Data Lab^a^. All these data sets are collected from the date on which the total number of cumulative daily cases for each of these countries crossed a count of 100. For Spain, we collected daily confirmed cases data from March 03 to June 13, 2020, whereas for Italy, it was collected from February 24 up to June 17, 2020. The univariate time series data set for Spain contains 103 observations and 115 observations for Italy. The datasets are divided into training and test set by setting a ratio of 2:1. We have used 2/3 of the data set for training the models, and the model performance is evaluated on the test data set containing 1/3 of the total data. In the following subsections, we discuss the necessary data pre-processing steps, followed by the development of the proposed integrated approach.

### 2.2. Data Pre-processing Stage

In the data pre-processing stage, we verify the presence of long-term memory, deterministic and stochastic components in the time series data sets of Italy and Spain. The long term memory property of a time series is measured using the Hurst exponent (HE). The value of HE lying between 0.5 and 1 proves that the series is sufficiently long. To calculate the HE for the given data sets, we use *‘pracma’* package in R statistical software. For daily cases data of Spain, the value of HE is 0.775 and 0.753 for Italy. Thus, it is confirmed that the data sets have long-term memory. We also plot the training and test data for Spain and Italy in Fig. 1, which confirms the presence of both the deterministic and stochastic behaviors in the COVID-19 daily cases data. This intrigued the need for an integrated deterministic-stochastic approach.

**Figure 1:**
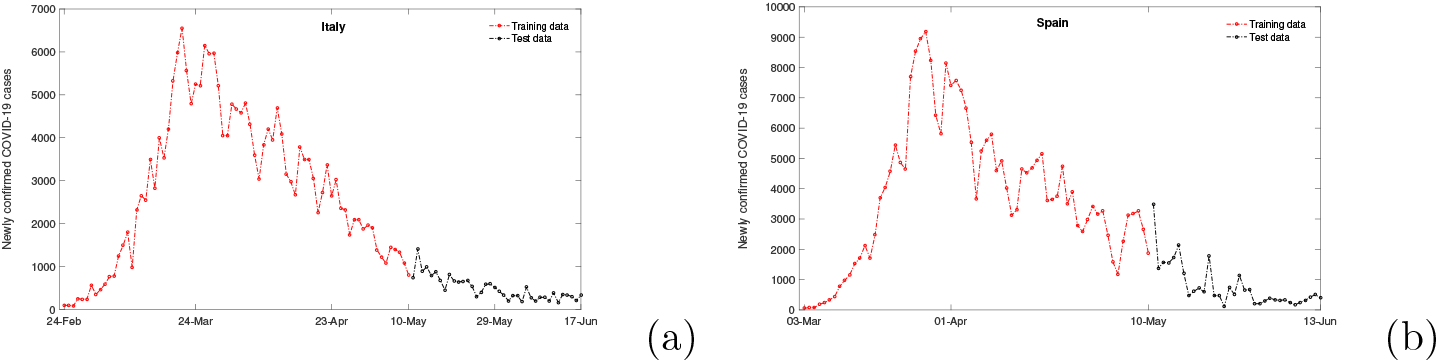
Plots of training and test data for (a) Italy and (b) Spain.

### 2.3. Proposed Integrated SIRCX-AR (ISA) Model

To find the long-term forecasts of the confirmed cases of COVID-19, we adopt an integrated deterministic-stochastic approach combining compartmental SIRCX and stochastic AR model. The proposed combined model blends two different modeling paradigms to exploit their individual benefits and overcome their shortcomings. In general, the proposed modeling process consists of three major steps:

- Modeling the deterministic components with an extended SIR (SIRCX) model;
- Evaluation of the SIRCX model residuals and finding the presence of uncertainties in error terms;
- Stochastic modeling of the left-out SIRCX residual values by the traditional AR model.

We assume that the daily COVID-19 cases *Q*(*t*) comprises both the deterministic (*D*(*t*)) and stochastic (*P* (*t*)) components as follows:

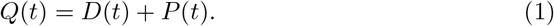

To predict the long-term trajectories of the COVID-19 cases, the deterministic model is initially used, followed by its residuals, which consist of some unexplained stochastic components (*P* (*t*)). This assumption holds for the COVID-19 data sets since the residual series produced by the mathematical model has no deterministic terms, as shown in Fig. 3. We can now estimate both *D*(*t*) and *P* (*t*) from the available daily cases data set. Let 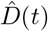 be the long-term forecasts based on the SIRCX model at time t, and *P* (*t*) represents the residuals containing the stochastic components at time t, obtained from the SIRCX model. Thus, we write

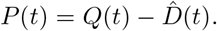

**Figure 2:**
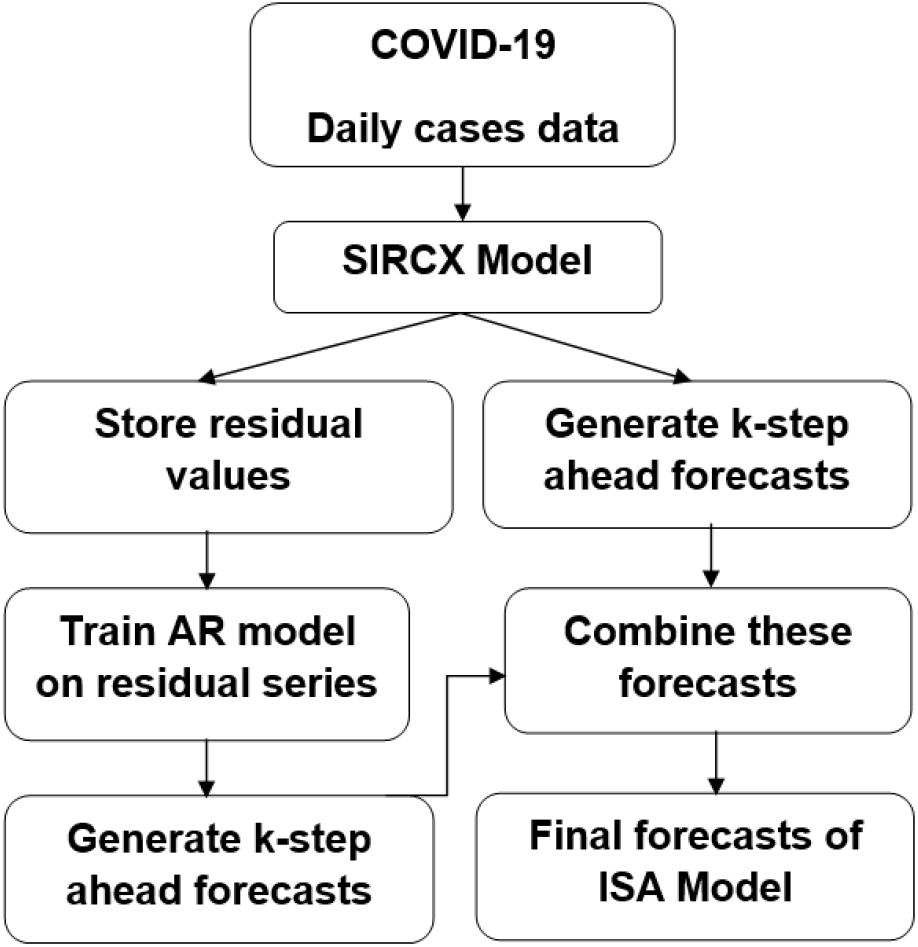
Flow diagram of the proposed Integrated SIRCX-AR (ISA) model

**Figure 3:**
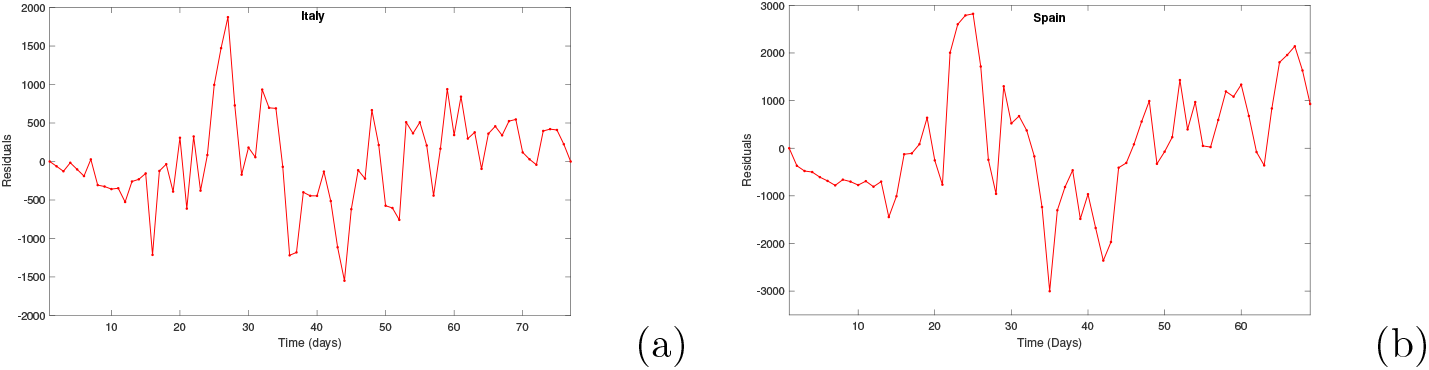
Plots of leftover ODE residuals for (a) Italy and (b) Spain.

These residuals are remodeled with an AR model, and predictions are obtained as 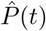. Therefore, we write the combined forecast as:

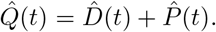

The proposed ISA model is a joint approach based on deterministic and stochastic methods. Figure 2 shows the systematic overview of the proposed integrated SIRCX-AR or simply ISA model. Below we discuss the constituent models (SIRCX and AR) used in the ISA model in detail.

#### Remark 1

The proposed ISA model combines two contrasting paradigms to forecast long-term trajectories of COVID-19 data. We choose two completely diverse models (SIRCX and AR) for hybridization, one from mathematical epidemiology literature and another from the area of traditional time series forecasting. The newly introduced integrated model will be practically useful for a better understanding of the dynamics of the complicated COVID-19 pandemic, as shown in Section 3.

#### 2.3.1. Modeling deterministic components with SIRCX Model

We propose an extension of a generalized SIR modeling, namely SIRCX model that reflects the known epidemiology of COVID-19, following [9]. When basic epidemiological parameters are largely uncertain, dynamical equation based compartmental models can often provide some insights on the long-term dynamics. Here, the total population is assumed to be constant, viz. we assume the birth rate and natural death rate have the same value (*µ*). We consider five disjoint compartments in the SIRCX model: susceptible *S*(*t*), infected *I*(*t*), recovered *R*(*t*), protected *C*(*t*) and isolated *X*(*t*). Protected people are those who take personal protective measures such as use of face mask. The susceptible individuals can become infected after a successful contact with an infected person. We also assume that isolated persons can not transmit the disease whereas the protected people exercise social distancing and can not become infected. The dynamics of COVID-19 are governed by the following system of equations:

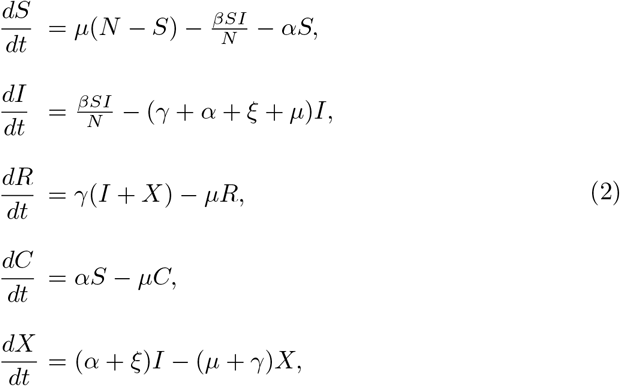

where *N* = *S* + *I* + *R* + *C* + *X*. The parameters of system (2) are the rate at which susceptible people adopt personal protection measures (*α*), the infection rate (*β*), the recovery rate (*γ*), the isolation rate (*ξ*) and the natural death and birth rate *µ* = (Life expectency)^*−*1^. The basic Reproduction number for SIRCX model is found to be

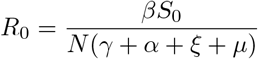

where *S*_0_ is the initial number of susceptibles in the system, as decribed below.

##### Proposition 2.1

*The system* (2) *is bounded in the region*

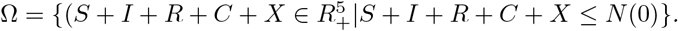

**Proof**. Adding all the compartments of system (2), we have

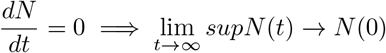

Hence the system (2) is bounded in the region Ω.

Further, we determine the condition for disease progression, when most of the people in the community are susceptible (*S*). In this kind of situation, *S* in the model (2) can be replaced by initial number of susceptibles, i.e., *S*(0) = *S*_0_. Therefore, the equation for infected people of the model (2) becomes

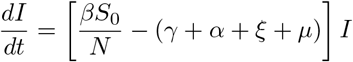

Therefore, we can obtain the solution to the differential equation as *I*(*t*) = *I*_0_*e*^*δt*^, where *I*_0_ is the initial number of infected people. Therefore, we have

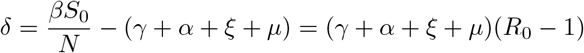

where, 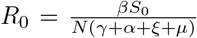 represents the basic reproduction number of the model (2). This threshold quantity is one of the most important parameter in the spread of an infectious disease in the community [13]. From the expression of *R*_0_, it can be noted that isolation rate and adoption rate of personal protection measures are inversely proportional to *R*_0_.

#### 2.3.2. Modeling stochastic components with AR model

Once the SIRCX model models the deterministic trend, we can now remodel the left-out uncertainties (error terms) with an AR model. AR model provides a parsimonious description of a (daily) stationary time-dependent stochastic process in terms of autoregressive terms. AR(*p*) denotes the AR model, where the parameter *p* is the order of the model. Since we removed the deterministic component of the series by a compartmental model and AR is applied to model the residual components of SIRCX. Thus no differencing and moving average terms are required in the model. AR(*p*) model can explain the momentum and mean reversion effects of the unexplained error terms that the deterministic model could not capture. AR model can be mathematically expressed as follows:

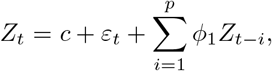

where *Z*_*t*_ denotes the actual value of the variable under consideration at time *t, ε*_*t*_ is the random error at time *t, c* is a constant and *φ*_*i*_ is the coefficients of the AR model. The ‘best’ fitted AR model is selected based on the Akaike’s information criteria (AIC) [14].

## 3. Experimental Analysis

In this section, we test the proposed methodology on the COVID-19 daily cases data sets for Italy and Spain. In the proposed ISA model, the deterministic trend in the series is first modeled using SIRCX model (2). To perform the data fitting, we used a simplex alogorithm-based function ‘*fminsearchbnd* ‘ (MatLab, R2016a) that minimizes the sum of squared errors (SSE) between simulated indicator

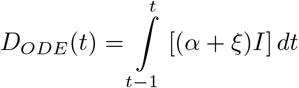

and reported data. Suppose we have *M* independent observations of daily COVID-19 cases, then we minimize 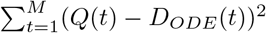, where *Q*(*t*) is daily reported COVID-19 cases.

The values of the fixed parameters, initials conditions, and estimated parameters are reported in Table 1. Using the fitted SIRCX model, we obtain the predicted values and two months ahead forecasts for COVID-19 cases of Italy and Spain.

**Table 1:** Parameters and initial values used for fitting SIRCX model

| Country | Parameter/ICs | Value | References |
| --- | --- | --- | --- |
| Italy | $\beta$ | 0.3745 | Estimated |
| | $\alpha$ | 0.0470 | Estimated |
| | $\xi$ | 0.01 | Estimated |
| | $\gamma$ | 0.0213 | [7] |
| | $\mu$ | $3.2616 \times 10^{-5}$ | [15] |
| | $N$ | 6,04,61,826 | [15] |
| | $I(0)$ | 2345 | Estimated |
| Spain | $\beta$ | 0.3468 | Estimated |
| | $\alpha$ | 0.0396 | Estimated |
| | $\xi$ | 0.01 | Estimated |
| | $\gamma$ | 0.057 | [16] |
| | $\mu$ | $3.2616 \times 10^{-5}$ | [15] |
| | $N$ | 4,67,54,778 | [15] |
| | $I(0)$ | 7912 | Estimated |

The leftover residuals obtained from the SIRCX model (also see Figure 3) are further modeled with an AR(*p*) model by using *‘forecast’* package in R statistical software. The order of AR(*p*) model is specified from the autocorrelation function plots (given in Figure 4). AR(5) was fitted to the residual series of Italy data having AIC = 1171.59 and log-likelihood value as -576.79. For Spain, AR(3) was fitted on the residual series with AIC=1168.08, and Log-likelihood value equals - 556.23. Using the ‘best’ fitted AR models, two months ahead mean forecasts of the residuals are generated for Italy and Spain along with the confidence intervals.

**Figure 4:**
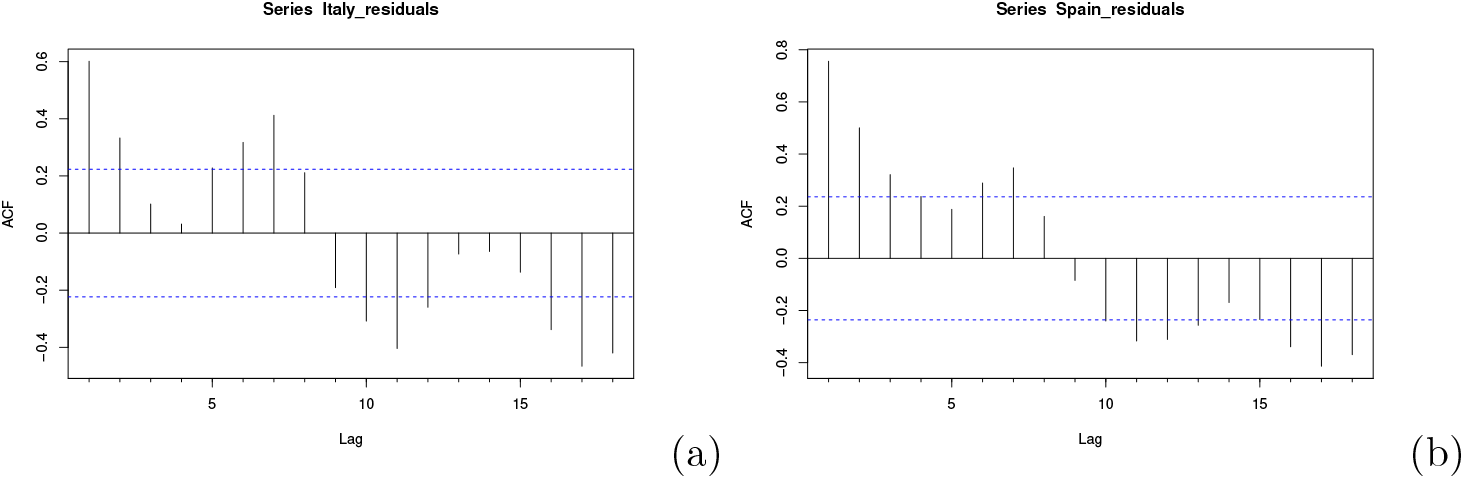
Plots of ACF values on ODE residuals of (a) Italy and (b) Spain.

Finally, the ODE generated forecasts and AR-based residual forecasts were added together to get the final forecasts. The ODE model predictions and ISA model forecasts are then compared using four widely used goodness-of-fit metrics, namely symmetric mean absolute percentage error (SMAPE), root mean squared error (RMSE) and mean absolute error (MAE). By convention, the models with lower values of goodness-of-fit metrics are capable of giving better forecasts. The formulae to evaluate these performance metrics are as follows:

1. 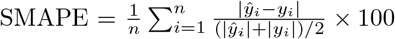;
2. 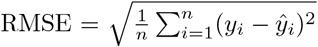;
3. 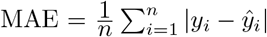;

where *y*_*i*_ are reported cases, *ŷ*_*i*_ are the predicted values and *n* denotes the total number of observations. The values of these metrics for Italy and Spain are reported in Table 2. From Table 2, we can conclude that the integrated SIRCX-AR (ISA) model outperformed the individual SIRCX model in both training and test COVID-19 data sets for Italy and Spain in significant margins irrespective of the performance measures.

**Table 2:**
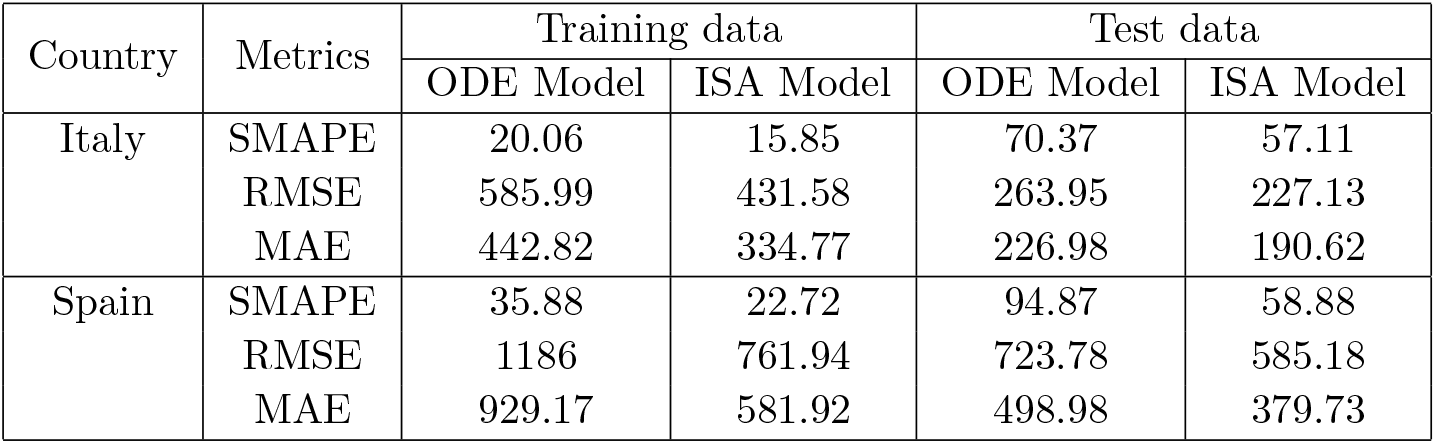
Performance metrics of SIRCX model and ISA model for Italy and Spain

We predicted two month ahead forecasts (long-term) for both Spain and Italy to understand the future outbreaks of the pandemic for Italy and Spain. The plots of actual vs. predicted values along with the long-term forecast values are given in Figure 5. Figure 5 depicts that the proposed ISA approach fitted the training data more adequately in comparison with the ODE-based SIRCX model. The bandwidth presented in Figure 5 shows slow decay in the number of future COVID-19 outbreaks in Italy and Spain. The predicted vs. actual forecasts suggest that the integrated approach is more accurate than merely relying on the compartmental model. The proposed ISA model assessed the asymmetric risks involved in the series way better than the individual epidemiological model. The proposed ISA model can easily be updated periodically (weekly) once more data becomes available.

**Figure 5:**
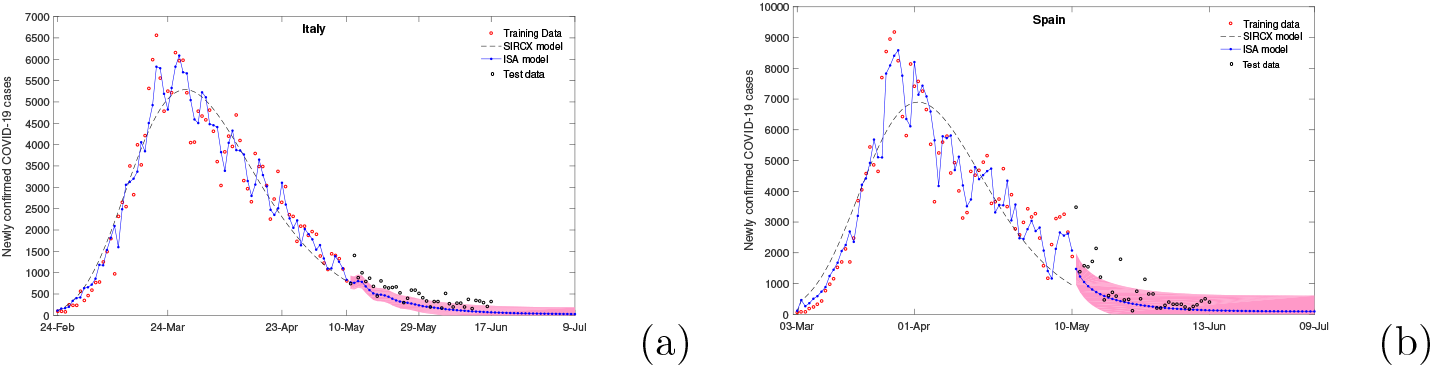
Fitting and long-term forecast of COVID-19 cases in (a) Italy and (b) Spain.

Using parameter values from Table 1, the basic reproduction numbers for Italy and Spain are found to be 4.78 and 3.25, respectively. These values indicate that COVID-19 spreads rapidly in a population unless containment strategies such as social distancing and effective isolation of infected individuals are implemented. From the long term trajectories, we estimated the expected number of cases in the next 30 days (May 11, 2020 – June 09, 2020) for the two countries. The estimated number of cases in Italy and Spain in the next 30 days is 10982 (6383 – 15582) and 13731 (3395 – 29013), respectively. Furthermore, the expected number of daily cases on 60^*th*^ day (July 09, 2020) for Italy and Spain is estimated to be 30 (0 – 183) and 92 (0 – 602), respectively. These numbers are very close to the actual number of COVID-19 cases that occurred in these two countries. Overall, these two countries have effectively controlled the burden of COVID-19 with effective control strategies.

## 4. Discussions

The global spread of pandemic COVID-19 poses a significant threat to the health-care systems of severely affected countries. The availability of limited data and inadequate epidemiological knowledge make the problem more challenging. Mathematical and statistical forecasting models always give insightful information about the future of COVID-19 spread in the community. In this study, we proposed an integrated deterministic-stochastic framework for predicting the long-term trajectories of COVID-19 cases in two profoundly affected countries, namely Italy and Spain. Combining two contrasting paradigms (deterministic and stochastic models) presented in this study is relatively new. We assumed that the time-dependent stochastic process consists of both the deterministic and stochastic components and examined it for the daily case data sets of COVID-19. The proposed ISA model outperformed the individual SIRCX model in terms of four performance metrics on both training and test data. Two month-ahead forecasts are provided for Italy and Spain. The estimated number of cases in Italy and Spain from May 11, 2020, to June 09, 2020, is 10982 (6383–15582) and 13731 (3395–29013), respectively. Furthermore, the expected number of daily cases on 60^*th*^ day (July 09, 2020) for Italy and Spain is estimated to be 30 (0–183) and 92 (0–602), respectively. These results indicate that the ISA model can effectively forecast the long-term trajectories of COVID-19 for test data sets. However, the current mitigation and control interventions should be maintained as there are high values of basic reproduction numbers for Italy and Spain (4.78 and 3.25, respectively) being observed to prevent any future outbreaks.

However, there are certain limitations of the proposed integrated framework. While formulating the ODE model we made some simplifying assumptions: (a) the birth rate and death rate of the population are constant, (b) we only consider homogeneous mixing of population, (c) the recovered people will get permanent immunity and (d) the isolated individuals can not transmit the disease and contained people can not get the infection. These assumptions can be relaxed in the future study. The proposed integrated deterministic-stochastic approach will be best-suited when the peak of the epidemic is passed. As a future scope of research, more complex statistical models instead of the AR model can be employed to model the leftover residuals of the ODE model. Though the model arrived from the analysis of COVID-19 data of Italy and Spain, the proposed ISA model will be well-versed for other profoundly affected countries and also for other similar epidemics.

## Data Availability

Available on request.

https://www.worldometers.info/coronavirus/

## Footnotes

a https://ourworldindata.org/coronavirus

